# Therapeutic potential of the genus *Bacteroides* in hypertension: an intervention of long-term intermittent fasting

**DOI:** 10.1101/2025.04.10.25325255

**Authors:** Junhong Su, Fanglin Li, Wenlian Bai, Zhongren Ma, Maikel P. Peppelenbosch

## Abstract

Intermittent fasting has shown promise in the management of hypertension, but the mechanistic explanation for this effect remains largely obscure. Studies in experimental animals suggest that intermittent fasting acts on hypertension by modifying the microbiome and particularly by increasing levels of intestinal Bacteroides. Human data, however, are lacking. Here we conducted a clinical trial [ChiCTR2000034646] to investigate the effects of 15-week intermittent fasting on individuals with hypertension. We observe that long-term intermittent fasting effectively counteracts high blood pressure, as demonstrated by significant reduction in systolic blood pressure (144±4.8 [S.E.M.] mmHg at baseline vs. 129±5.6 mmHg at 15 weeks, p=0.004) and diastolic blood pressure (94±5.2 mmHg vs. 79±2.9 mmHg, p=0.005), as well as serum uric acid (410 ± 38 mmol/L at baseline vs. 307 ± 5.5 mmol/L at 15 weeks, p=0.032), which is a strong risk marker for hypertension. Importantly, this effect is associated with significant remodeling of the fecal microbiome (p=0.041). Mirroring earlier data in experimental rodents, we observe a strong inverse correlation between levels of genus Bacteroides and blood pressure (R=-0.608, p=0.04). Our results strongly support the notion that the genus Bacteroides is a major determinant of blood pressure in hypertensive individuals.

**Importance:** While intermittent fasting is generally recognized to be beneficial for patients with high blood pressure, the mechanistic basis for this effect is not resolved. Based on animal data, however, a role of the microbiome and especially the genus Bacteroides has been suggested. Thus prompted, we conducted a clinical trial in hypertensive individuals to link the fecal microbiome to changes in blood pressure. We observed strong correlations between improved blood pressure and fecal levels of the genus Bacteroides. In conjunction with the body of contemporary biomedical literature our data suggest that the effect of intermittent fasting on blood pressure is mediated through Bacteroides opening a novel avenue for rational treatment of this condition.

## OBSERVATION

Hypertension is a significant public health issue worldwide, responsible for approximately 54.5% of ischemic heart disease and 50.0-58.3% of stroke deaths each year. Poor lifestyle choices, including obesity and an unhealthy diet, are driving factors for hypertension^1^. Although it is well established that the pathogenesis of hypertension is primarily characterized by decreased vasodilation and increased blood volume, recent studies indicate that dysbiosis of the gut microbiome is potentially an important factor in its pathogenesis, potentially driving changes in vasotension^2^. In this context, it is important to note that drugs used to manage hypertension can also affect the microbiome, aggravating the dysbiosis and thus counteracting efficacy^3, 4^. Identifying the elements in the microbiome that interact with hypertensive disease would, therefore, be valuable for refining strategies to manage hypertensive patients effectively.

More specifically, the depletion of short-chain fatty acid (SCFA)-producing bacteria is a critical feature of a dysbiotic gut microbiota in hypertension^2^. Notably, Bacteroides, which is a major producer of intestinal propionate and a significant contributor to the enterotypes in healthy individuals, is substantially reduced in hypertension^5-7^. Building on this, research demonstrates that enhancing the levels of intestinal Bacteroides in animals can help reduce blood pressure through mechanisms involving the regulation of intestinal steroid hormone levels^5^ and the restoration of bile acid metabolism, suggesting that enhancing the levels of Bacteroides may hold promise in addressing clinical hypertension. However, to date, no clinical trials have reported associations between altered levels of Bacteroides and the clinical course of hypertension. This lack of data substantially impedes progress in the field.

Our previous work demonstrated that one month of 16-hour intermittent fasting significantly increases the level of intestinal SCFAs-producing bacteria in healthy volunteers^8^. Hence, we hypothesized that intermittent fasting may improve blood pressure by increasing intestinal SCFA producers. To directly investigate this notion, we designed a study in which calorie intake was restricted from 8:00 am to 14:00 pm continuously for 15 weeks (15wksIF) in hypertensive individuals (**Figure 1A**). Patients were only allowed to consume no more than 3 bananas or 3 apples or the combination when they felt very hungry before going to bed. Patient baseline information is shown in **Supplementary Table 1**.

**Figure 1.**
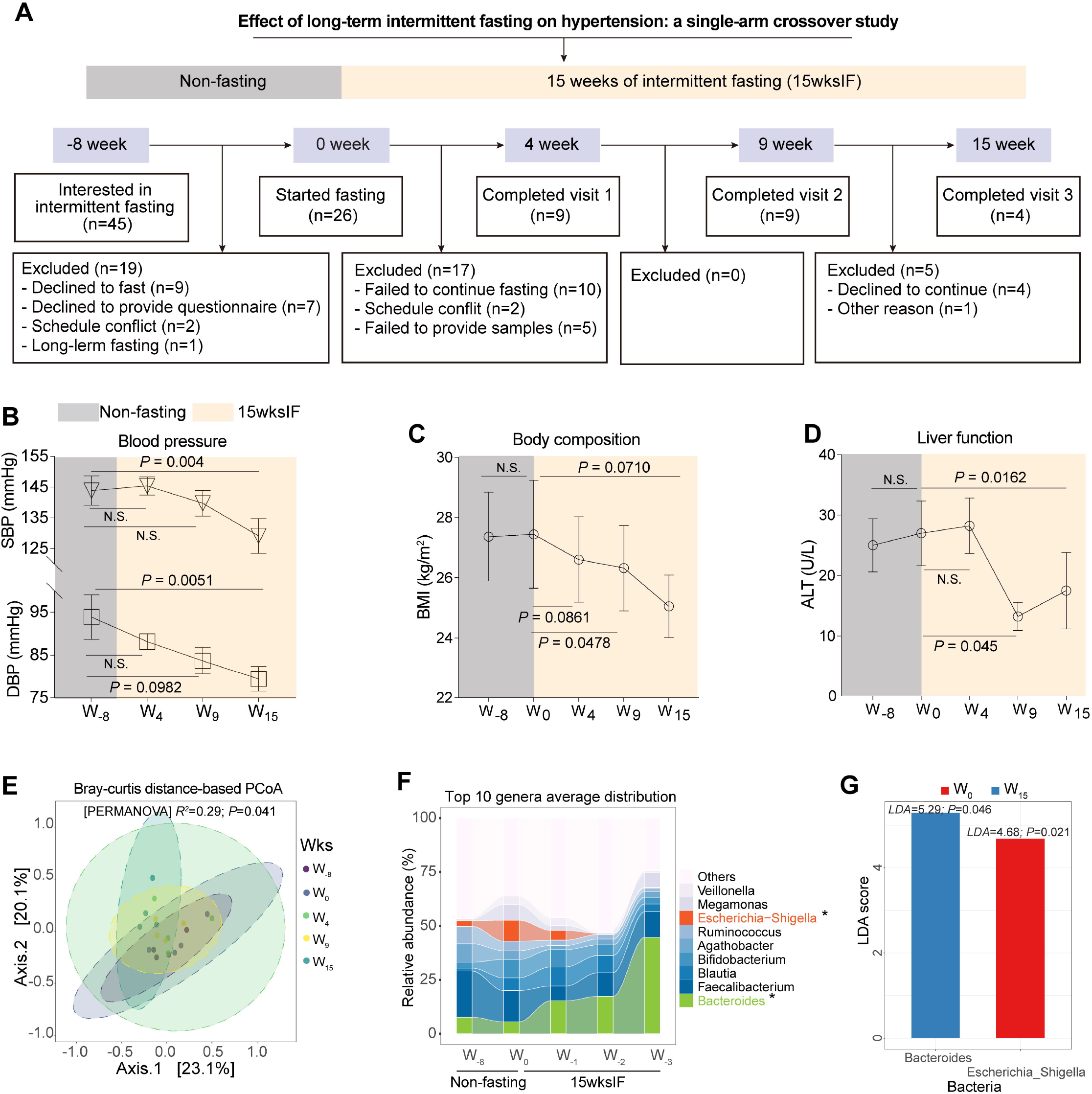
Effects of intermittent fasting on blood pressure and the gut microbiome in hypertensive patients. (**A**) Study flowchart. The change in blood pressure (**B**), an independent risk factor BMI (**C**) and liver function indicator ALT (**D**). Data are expressed as mean ± S.E.M. P values are calculated using paired student’s *t*-test (one-tailed). (**E**) The overall microbiome composition in patients was represented visually using principal coordinate analysis (PCoA) of genus-level relative abundance. The samples that are clustered as individual centroids are from different time point denoted by different colors. P values were calculated using PERMANOVA test. (**F**) Sankey plots showing the longitudinal change of genus level abundance during intermittent fasting. P values are calculated using paired student’s *t*-test (one-tailed). (**G**) Taxa that differed at baseline at week 0 versus end of fasting at week 15 are depicted and visualized in bars using LEfSe with a log LDA (linear discriminant analysis) score above 4.0. Blue bar represents taxa where increases in its relative abundance or prevalence associated with 15wksIF, while red bar means taxa decreased during 15wksIF.

At the end of 15 wksIF, participants showed substantial improvement in both systolic blood pressure (SBP) (144±4.8 [S.E.M.] mmHg at baseline vs. 129±5.6 mmHg at 15 weeks, p=0.004) and diastolic blood pressure (DBP) (94±5.2 mmHg vs. 79±2.9 mmHg, p=0.005) (**Figure 1B**). Improvements in clinical hypertension were further objectified by a significant reduction in circulating uric acid (410 ± 38 mmol/L at baseline vs. 307 ± 5.5 mmol/L at 15 weeks, p = 0.032; **Supplementary Figure 1B**), a marker for hypertension^9^, and indirectly validated by improvements in two independent risk factors: BMI and liver function (**Figure 1C and 1D**). However, changes in blood pressure and associated clinical parameters were not significant after 4 weeks of fasting, indicating that prolonged fasting is a necessary determinant for the clinical success of intermittent fasting in lowering blood pressure.

To investigate the notion that changes in the gut microbiome-especially with regard to the levels of the genus Bacteroides-might be instrumental for mediating the effects of fasting on blood pressure^10^, the gut microbiome was examined before fasting (week-8) and during fasting (week 0, 4,9,15) using fecal sampling and 16S sequencing (**details in Supplementary Materials**). Employing the data obtained, compositional analysis using Bray-Curtis distance-based principal coordinate analysis revealed significant changes in the intestinal microbiome of hypertensive participants after 15wksIF (p = 0.041 by PERMANOVA test) (**Figure 1C**), demonstrating substantial shifts in the levels of specific high-abundance taxa.

More specifically, Sankey plot showed that the Firmicutes (71.6 ± 5.92 [S.E.M.] %) and Proteobacteria (11.93 ± 3.83%), which were the dominating phyla at baseline, decreased to 46.67 ± 11.39% (p=0.0122) and 1.74 + 0.64% (p=0.0401), respectively, following 15wksIF (**Supplementary Figure 1A**). In contrast, Bacteroidetes increased from 5.58 ± 1.58% to 44.66 ± 11.27 % (p=0.0241), becoming the most abundant phylum. This increase in Bacteroidetes could be further attributed to a significant rise in sequences from the genus *Bacteroides* (**Figure 1E**), which were 8.01 times higher than before fasting and included primarily *B. stercoris, B. uniformis* and *B. vulgatus*). The change of this genus was validated by Lefse analysis **(Figure 1F)** and hierarchical heat tree analysis (**Supplementary Figure 1B**). Noticeably, *Bacteroides* levels were inversely correlated with systolic blood pressure (R=-0.608, P=0.04), suggesting a causal connection between this bacterium and the control of blood pressure (**Supplementary Figure 1C**), supporting the findings in animals by others that changes in Bacteroides abundance relate to hypertension^5^.

Interestingly, the administration of living *Bacteroides* to experimental animals was found to reduce intestinal endotoxin levels by decreasing the size of *E. Shigella* compartment^11^, which aligns well with the notion that the reduction in *E. Shigella* in our patients reflects increased size of the *Bacteroides* compartment, in turn suppressing *Escherichia* (**Supplementary Figure 1D**; Spearman’s ρ=-0.517, p=0.0195). Additionally, intake of rice and wheat-based food during fasting might also influence microbiome regulation (**Supplementary Figure 1E**), which warrants further investigation.

To rule out the possibility that patients who maintained intermittent fasting throughout the entire study period might have a different baseline microbiome, which could obscure the effects of fasting, we compared the gut microbiome between non-completers and completers at baseline (0 weeks). No significant differences were observed in microbiome diversity (P=0.4209) (**Supplementary Figure 1F**), microbiome composition (P=0.304 by ANOSIM test) (**Supplementary Figure 1G**) or the relative abundance of the *Bacteroides* (P=0.2622) (**Supplementary Figure 1H**). Hence the most straightforward explanation is that a strong negative correlation exists between blood pressure and levels of intestinal *Bacteroides*.

## Conclusion

We observed that 15wksIF is effective in improving hypertension by increasing intestinal *Bacteroides* levels, although patient’s motivation and capacity to maintain such a regimen may be limited, particularly in males^12^. Furthermore, this study provides a promising perspective that Bacteroides may serve as a novel bacterial candidate for restoring an imbalanced gut microbiome and improving blood pressure. This insight opens new avenues for the rational design of beneficial Bacteroides-based therapeutic strategies to address this condition, with potential to complement current treatment that carry side effect and exacerbate dysbiosis of the gut microbiome.

## Acknowledgements

The authors would like to acknowledge Yueying Wang and Yuxin Su for their assistance in sample collection and questionary survey. The authors are grateful to all patients who contributed to this work. This study was supported in part by grant (KICH2.V4P.22.015) of the Dutch Organization for Scientific Research and the Dutch Cancer Foundation.

## Data availability

The sequence data have been deposited with links to BioProject accession No. PRJNA1118570 in the DNA Data Bank of BioProject database (https://www.ncbi.nlm.nih.gov/bioproject/PRJNA1118570). Source data of figures is included in the supporting files.

