## Supplemental information for "Therapeutic potential of the genus *Bacteroides* in hypertension: an intervention of long-term intermittent fasting"

### Supplementary information

**Supplementary Methods**

**Patients and execution of intermittent fasting**

We prospectively recruited hypertensive patients who expressed the intention to observe 16:8 intermittent fasting for up to 15 weeks (15wkIF). The fasting period of each day was from 14:00 pm to 8:00 am next day. Patient recruitment was limited to database list in People's Hospital of Linxia Hui Autonomous Prefecture, where those who have been diagnosed with hypertension in past years was called for their interest in participating of the study. The diagnostic criteria for hypertension are SBP > 140 mmHg and/or DBP> 90 mmHg without antihypertensive medications. Exclusion criteria were self-reported disease cancer, cirrhosis, arthritis, nephritis, chronic heart failure, tuberculosis, anemia, chronic respiratory disorders, or any gastrointestinal disorders. In this study, we designed a sing-arm within cross-over trail, in which selected patients were first requested to eat ad libitum for eight weeks, followed by 15 weeks of intermittent fasting. The stool of each patient was collected before fasting (week-8) and during fasting (week 0, 4,9,15). Finally, 26 patients who met the study criteria were included. During the intervention, seventeen patients dropped out at the secondary visit and three at the fourth visit. The remaining patients, all female, complemented 15 weeks of fasting according to the study protocol. The stool of each patient was collected before fasting (week-8) and during fasting (week 0, 4,9,15). To avoid environmental contamination, all volunteers were kindly requested to 1) wash their hands before sampling their own stool, 2) place a clean toilet paper to catch the stool and 3) collect the stool in a clean, screw-top container by using the spoon that came with the container. Following collection, stool samples were transported on ice to the laboratory and stored at –80°C until further processing.

**Ethics approval and consent to participate**

The protocol of this study was approved by the medical ethical committee of Northwest Minzu University (XBMZ-YX-2,016,001). This study is registered at www.chictr.org.cn (ChiCTR2000034646). All volunteers provided written informed consent prior to the study.

**Blood pressure measurement**

Blood pressure was measured manually by a table-top blood pressure monitor for all time points except the last one, at which blood pressure was measured by a 24-hours ambulatory blood pressure monitors and its average value was used for statistical comparison. Data on blood pressure at week 0 was missing.

**Estimation of dietary intake**

To assess food intake associated with 15 weeks of intermittent fasting in the present cohort, the change in different food intake over the whole course of the study was estimated via modified food frequency questionnaire (FFQs), identical to the methodology described elsewhere (1). Included patients reported their food intake frequency through a face-to-face interview at each visit. Briefly, the following 5 frequency categories were designed for performing FFQ: never eaten, eating daily, eating weekly, eating monthly, and eating yearly. This FFQ enables us to accurately category different food items based on their popularity during each month of the study. We did not include food types that were consumed on a yearly basis.

**Outcome**

The primary outcome was the change in gut microbiota composition after 15wksIF intervention compared with baseline. To this end, gut microbiota were analyzed for diversity, composition, and taxonomic abundance. Secondary outcome variables were changes in body composition, blood parameters, and food intake.

**Characterization and analysis of the gut microbiome**

Fecal DNA was extracted from the stool sample using by a modification of the cetyltrimethylammonium bromide method as described elsewhere in detail (2). DNA quality was monitored on 1% agarose gels. The quality of the library was assessed using the Qubit@ 2.0 Fluorometer (Thermo Scientific). Next-generation sequencing of the V3–4 region of the bacterial 16S ribosomal RNA gene was performed by Illumina MiSeq platform. Data processing and analysis were performed according to routine procedures, as described elsewhere in detail (1, 3). Briefly, by using DATA2 (incorporated in QIIME2) (4), paired-end raw reads (250 bp) were denoised, demultiplexed, trimmed, of chimera removal and finally merged so as to construct an amplicon sequence variable (ASV) table. Representative sequences were classified against the SILVA (v138) reference database for taxonomy classification. The gut microbiome diversity was estimated by calculating the Shannon and Evenness using alpha-diversity.py in QIIME2 (5). The beta-diversity was done with Principal coordinate analysis (PCoA) using the R packages vegan (version 2.6.4). To further evaluate if the overall changes in the gut microbiome composition during intermittent fasting is significant, analysis of similarities (ANOSIM) test was performed using the R packages vegan (version 2.6.4) with 999 permutations. Taxa with significantly differential abundance between groups were visualized using heat trees with a Wilcoxon p-value test adjusted for multiple comparisons on the proportional microbiome data (6). To identify bacterial taxa whose sequences were differentially abundant between different time points, linear discriminant analysis (LDA) coupled with effect size measurements (LEfSe) was applied with the significance level of 0.05 and the logarithmic LDA score threshold equal to 4.5.

**Statistical analysis**

Statistical analyses were performed using R software, version 4.2.0, and Prism version 7.0 (GraphPad Software). The significance of differences between 2 groups was evaluated using the 1-tailed paired Student's t test. To discover the strength and direction of a link between 2 parameters, Pearson's rank correlation coefficient was calculated using the rcorr function from the Hmisc R package. The change of semi-quantified food intake was estimated using the Friedman test with uncorrected Dunn's multiple tests. Results were expressed as mean ± SEM. For all tests, a value of $P<0.05$ was considered to indicate statistical significance.

**Supplementary Figure 1.** Intermittent fasting on the gut microbiome in hypertension.

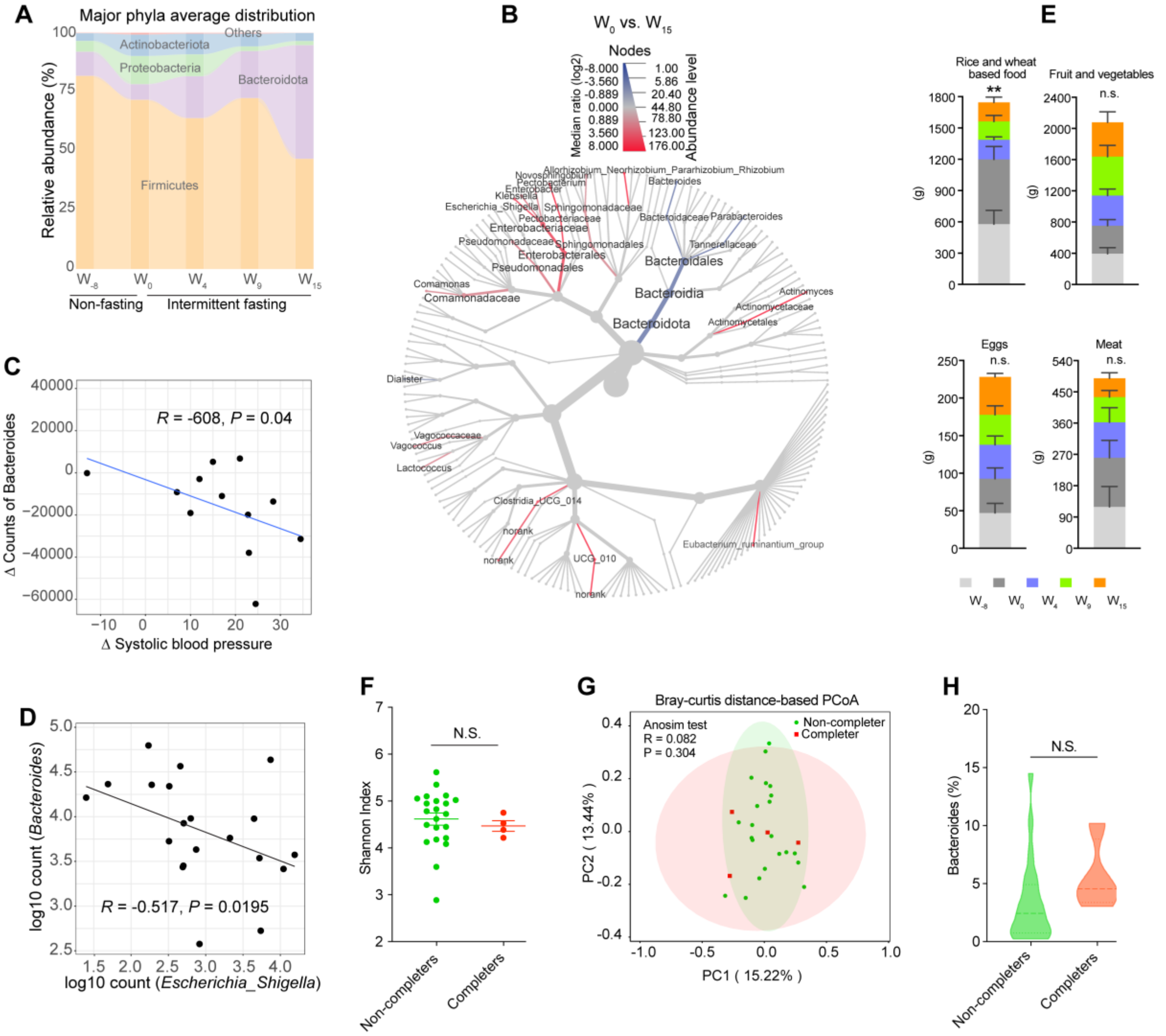


(A) Sankey plots showing the longitudinal change of phylum level abundance during intermittent fasting. P values are calculated using paired student's t-test (one-tailed). (B) Hierarchical differential heat tree depicting differently abundant taxa between two groups based on fasting durations (baseline at week 0 versus end of fasting at week 15). The node size and the color intensity reflect the species abundance and the log2 median proportion between the two groups respectively. (C) Correlation ecoefficiency between microbial biomarkers and blood pressure. Pearson's ρ and P value of the correlations are indicated. (D) Log-transformed counts of correlation (Pearson's ρ) between Bacteroides and Escherichia Shigella. (E) four most popular food categories were quantified according to a modified food questionnaire. Data represents mean ± S.E.M. Statistical analysis was performed by using the Friedman test with uncorrected Dunn's multiple tests. (F) The alpha diversity of gut microbiota between patients who dropped off (non-completer) and who completed the study (completer) was assessed by calculating the Shannon index. Data are expressed as mean ± S.E.M. P values are calculated using unpaired student's t-test (two-tailed). (G) The overall microbiome composition in non-completers and completers was not significantly different before intermittent fasting (p = 0.304), as represented visually using principal coordinate analysis (PCoA) of genus-level relative abundance. The samples that are clustered as individual centroids are from different time point denoted by different colors. P values were calculated using Anosim test with a condition of 999 permutations. (H) Relative absolute of Bacterodies at baseline was compared. No significant difference was observed ($p=0.2622$). Data are expressed as mean $\pm$ S.E.M. Statistical analysis was performed by using unpaired student's t-test (two-tailed).

**Supplementary Table 1.** Data of hypertensive patients before 15 weeks of fasting

| Variable | Non-completer (N=22) | Completer (N = 4) | P value |
|---|---|---|---|
| Age (yr) | 52.5 ± 3.57 | 51.5 ± 1.43 | 0.7882 |
| Sex female, *n* | 11 (50.00) | 4 (100) | 0.1134 |
| Smoking, *n* | 6 (27.27) | 0 (0.00) | 0.5425 |
| Alcohol, *n* | 6 (27.27) | 0 (0.00) | 0.5425 |
| Duration of disease (yr) | 7.10 ± 0.81 | 12.38 ± 6.61 | 0.1106 |
| Medication treatment, *n* | 22 (100) | 3 (75.00) | 0.7503 |
| Type 2 diabetes, *n* | 5 (22.72) | 2 (50.00) | 0.2580 |
| Vascular stent, *n* | 2 (9.09) | 1 (25.00) | 0.3596 |
| Sleep-wake cycle | | | |
| Bedtime | 10.41 ± 0.19 | 11.0 ± 0.41 | 0.2374 |
| Wake-up time | 6.41 ± 0.20 | 6.00 ± 0.41 | 0.4324 |
| Heavy activities | | | |
| Running, *n* | 1 (4.55) | 0 (0.00) | 0.6637 |
| Cycling, *n* | 3 (13.64) | 1 (25.00) | 0.5623 |
| Swimming, *n* | 0 (0.00) | 0 (0.00) | 1 |
| Climbing, *n* | 0 (0.00) | 0 (0.00) | 1 |
| Ball sport, *n* | 1 (4.55) | 0 (0.00) | 0.6637 |
| Others, *n* | 3 (13.64) | 1 (25.00) | 0.5623 |
| Body composition | | | |
| Weight, kg | 70.48 ± 3.45 | 73.75 ± 4.61 | 0.7005 |
| BMI | 25.43 ± 0.92 | 27.45 ± 1.79 | 0.3881 |
| Blood parameters | | | |
| Glucose, mmol/L | 5.47 ± 0.29 | 6.45 ± 0.72 | 0.1979 |
| HbA1c, % | 6.20 ± 0.27 | 6.60 ± 0.59 | 0.5242 |
| GGT, U/L | 29.14 ± 5.67 | 21.75 ± 3.35 | 0.5926 |
| AST, U/L | 20.86 ± 1.61 | 18.00 ± 2.04 | 0.4722 |
| ALT, U/L | 33.86 ± 3.28 | 27.00 ± 5.37 | 0.4043 |
| AST/ALT | 0.66 ± 0.03 | 0.73 ± 0.12 | 0.4408 |
| TCHOL, mmol/L | 4.30 ± 0.15 | 5.12 ± 0.37 | 0.0452 |
| HDL, mmol/L | 2.18 ± 0.19 | 3.12 ± 0.67 | 0.0807 |
| LDL, mmol/L | 3.41 ± 0.16 | 4.07 ± 0.23 | 0.1013 |
| Triglycerides, mmol/L | 3.36 ± 0.61 | 3.50 ± 1.47 | 0.9295 |
| Uric acids, µmol/L | 368.1 ± 16.36 | 409.8 ± 38.00 | 0.3272 |
| Blood pressure, mmHg | | | |
| Systolic | 124.1 ± 2.85 | 156.8 ± 6.24 | 0.0001 |
| Diastolic | 83.33 ± 2.33 | 99.75 ± 7.08 | 0.0132 |

**Note:** Numeric data was expressed as mean ± S.E.M. Categoric data are expressed as n (%). Significance between groups was estimated using two-sided un-paired student *t* test for numeric data or Fisher's exact test for categoric data. HbA1C, glycated hemoglobin; GGT, γ-glutamyltransferase; ALT, alanine aminotransferase; AST, aspartate aminotransferase; TCHOL, total cholesterol; HDL, high-density lipoprotein; LDL, low-density lipoprotein.
